# Daily EEG reveals stage-specific alpha power and functional connectivity modulation across five days of tACS in major depressive disorder

**DOI:** 10.64898/2026.03.17.26348546

**Authors:** Athena Stein, Tobias Schwippel, Francesca Pupillo, Hadden LaGarde, Mengsen Zhang, David Rubinow, Flavio Frohlich

## Abstract

**Background:** Major depressive disorder (MDD) is characterized by altered frontal alpha oscillations. Transcranial alternating current stimulation (tACS) can normalize aberrant oscillations in MDD, yet the daily dynamics of tACS target engagement of alpha oscillations in depression remain unclear.

**Methods:** In a double-blind randomized controlled trial (NCT03994081), 20 participants with MDD received verum or sham 10 Hz tACS (40 min/day, 5 days) targeted to left and right dorsolateral prefrontal cortex (F3/F4). High-density EEG was collected pre/post-stimulation each day to quantify within-session and cumulative changes in alpha power and functional connectivity (wPLI).

**Results:** Verum stimulation produced late-emerging, session-specific alpha power decreases compared to sham, with robust day (D)4 post-pre reductions at both IAF and 10 Hz across frontal and parietal regions (t=-2.42 to -3.82, p<0.05; parietal t=-3.82, pFDR<0.05). Whole-brain topographical analysis confirmed a distinct condition x D4 effect at left prefrontal cortex (t=2.9, pFWE<0.05, cluster permutation). Connectivity changes emerged earlier and more transiently, with D2 bilateral frontal wPLI reductions (t=-2.53, p<0.05). Cumulative analyses (change from D1) showed significant wPLI decreases on D2 and D3 (t=-2.65 and t=-2.46; p<0.05). Exploratory clinical correlations showed that the D4 IAF power decrease was associated with increased reward sensitivity (spearman rho= -0.6, p<0.05, cluster-corrected).

**Conclusions:** Alpha-tACS produced a temporally distinct neural response: an early, transient decrease in functional connectivity on D2, which may have driven a later suppression of left prefrontal alpha power on D4, correlated with clinical and behavioral improvements. These results delineate target engagement and validation mechanisms in a multi-day tACS trial, supporting optimized dosing in future tACS interventions.

## Introduction

Major depressive disorder (MDD) is a highly prevalent and disabling condition worldwide (1), and a substantial proportion of patients do not achieve full remission with existing pharmacological and psychotherapeutic treatments (2). Advances in circuit-targeted neuromodulation have opened a complementary, mechanistically grounded pathway for intervention. Among these, transcranial alternating current stimulation (tACS) offers a way to directly interact with abnormal neural oscillations that are increasingly implicated in the pathophysiology of MDD (3–6).

The alpha oscillation (8-12 Hz) has emerged as a compelling neurophysiological target for tACS in MDD, supported by a growing body of clinical trial evidence (3,6–11). Alpha is a prominent resting-state oscillation that is implicated in local inhibitory control as well as long-range network coordination, particularly within fronto-parietal and default mode network (DMN) circuits (12,13). Elevated alpha power, especially in frontal and fronto-central cortical regions, has been reported in MDD (14,15), and is suggested to relate to alterations in prefrontal excitation-inhibition (E/I) balance (16), disrupted network integration (17–20), and anhedonia-related impairments in reward processing (10,21,22). The left dorsolateral prefrontal cortex (dlPFC) is a key node in these circuits, tightly embedded within fronto-cingulo-striatal pathways that support cognitive control and affect regulation (23). In MDD, left dlPFC activity is strongly functionally coupled with the anterior cingulate cortex, demonstrated by both observational work (24) and transcranial magnetic stimulation trials (25,26). Together, these findings position alpha oscillations in the left prefrontal area as a mechanistically grounded target for interventions aiming to normalize pathological network states in depression, such as tACS.

tACS applies low-intensity oscillatory electric fields through the scalp to modulate endogenous cortical oscillations in a frequency specific manner (4,27,28). When applied at, or near, an individual’s intrinsic alpha frequency (IAF), tACS can entrain ongoing oscillations, thereby modulating alpha power and phase synchrony to affect both oscillations and functional connectivity (4,28–30), likely via a combination of short-term entrainment and longer-lasting synaptic plasticity (31). Recent developments in the therapeutic application of tACS include closed-loop protocols that personalize stimulation to IAF and adapt to ongoing neural activity (6), reflecting a move toward more precise, mechanism-based dosing strategies.

Clinical trials from our group have demonstrated that five days of alpha-tACS significantly reduces left fronto-central alpha power and improves depressive symptoms (3). Most recently, a pilot open label trial of closed-loop IAF stimulation demonstrated 80% remission after five consecutive sessions, with clinical improvement tied to a pronounced decrease in alpha power on Day 4 (6); although using a single channel EEG montage with poor spatial resolution. In contrast, the previous randomized trials recorded EEG only on the first and last stimulation days, meaning that the dynamics of brain activity during the treatment week remain largely undefined (32–34). Given the high participant and trial personnel burden of five consecutive days of stimulation, it is important to understand whether this truly represents the optimal dosing schedule.

In this study, we used high-density EEG (HD-EEG) to track daily neural dynamics during a five-day, sham-controlled 10Hz tACS protocol in MDD. HD-EEG was recorded immediately before and after each stimulation session, allowing characterization of immediate and cumulative changes in alpha power, individual alpha frequency, and functional connectivity in frontal and fronto-parietal networks. We also examined the stability and mismatch of IAF relative to the fixed 10 Hz stimulation across days, and tested whether day-specific neural changes, particularly in the prefrontal region, were related to changes in depressive symptoms and reward-related functioning. By moving beyond baseline-versus-endpoint contrasts, this work aims to track the neural dynamics of target engagement across the stimulation week and determine which of these markers, if any, track clinical improvement. To our knowledge, this is the first study to investigate the daily trajectory of neurophysiology in a tACS treatment trial for depression, providing important information about the time course of target and/or therapeutic effects.

## Methods and Materials

### Study design and outcomes

This was a randomized, double-blinded, sham-controlled clinical trial using a between-subjects design (ClinicalTrials.gov NCT03994081) conducted at the Carolina Center for the Neurostimulation, University of North Carolina at Chapel Hill. All participants provided informed consent, and research was conducted in accordance with the declaration of Helsinki. The University of North Carolina at Chapel Hill Institutional Review Board and Office of Human Research Ethics provided ethical approval for this study.

Participants first completed screening and eligibility confirmation procedures before attending our center for five consecutive daily sessions of verum or sham (placebo) 10Hz tACS. Clinician-administered and self-report clinical assessments of depression symptoms, mood, and quality of life were collected at Day (D)1, D5, and at a two-week post-stimulation follow-up (FUP). HD-EEG was recorded immediately before and after stimulation on each of the five days. Previous work has examined clinical and neurophysiological changes between D1, D5, and FUP (35). The current project aimed to more deeply characterize neurophysiological changes during tACS administration using HD-EEG. The primary aim was to investigate daily alpha power changes, including both immediate within-day and cumulative change from baseline. Secondary aims included investigating daily within-day and cumulative functional connectivity change, as well as the association of power/wPLI with clinical symptom changes.

### Participants

In this trial, we enrolled n=20 adults between the ages of 18-70 years with DSM-V diagnosed unipolar, non-psychotic MDD (Fig. S2). All participants had a Hamilton Depression Rating Scale (HDRS) score >8 and had low suicide risk, confirmed using the Columbia Suicide Severity Rating Scale (C-SSRS). Exclusion criteria included a diagnosis of moderate or severe alcohol/substance use disorder within the last 12 months, lifetime bipolar or psychotic disorder, eating disorder (within the past 6 months), obsessive-compulsive disorder (lifetime), post-traumatic stress disorder (within the last 6 months), as well as a change in antidepressant medication within the last 4 weeks, and/or on current antidepressant medication taken for less than 4 weeks.

### Sample size, randomization and blinding

The sample size (n=10 per group) was calculated based on a previous pilot trial conducted by our group, which reported a significant reduction in alpha power after five days of 10Hz tACS in a group size of n=9 (3). Randomization was conducted using a computer-generated list created by a researcher with no participant interaction (M.Z.). Participants and intervention staff were blinded to study group assignment.

### Clinical assessments

The HDRS-17 (36) was administered by a board-certified psychiatrist (T.S.) to assess depression severity at D1, D5, and FUP. A variety of self-report assessments were also conducted at the same timepoints: Beck Depression Inventory (BDI)-II (37), Snaith-Hamilton Pleasure Scale (SHAPS) (38), Behavioral Inhibition and Behavioral Activation Self Report Scales (BIS/BAS) (39). Due to a technical error, auxiliary self-reports are missing in participants 1-5 (n=3 verum, n=2 sham)

### Experimental procedures

#### Transcranial alternating current stimulation (tACS)

tACS was administered at 10 Hz (alpha) at 2mA zero-to-peak using the XSCITE device (Pulvinar Neuro, Durham, NC). Three conductive carbon rubber stimulation electrodes were positioned at scalp locations F3, F4 (5×5 cm; 1mA zero-to-peak at each location; in-phase), and Cz (5×7 cm; 2mA, anti-phase), according to the 10-20 EEG measurement system. Stimulation was applied for 40 minutes, including a ramp-up and ramp-down of 20 seconds. Identical procedures were used to administer active sham stimulation, except stimulation was ramped up for 20 seconds, maintained for 40 seconds, and then ramped down for 20 seconds (total 1 minute 20 seconds of stimulation), to mimic the sensations of stimulation. During tACS administration, participants viewed a relaxing video of fish swimming on a coral reef with flickering sunlight (Reefscapes; Undersea Productions, Queensland, Australia), designed to standardize brain state and to mask phosphenes to improve blinding (3).

#### EEG acquisition and preprocessing

Resting-state eyes-open HD-EEG was recorded at 1000 Hz with a 128-channel system using a NetAmps 410 amplifier (Electrical Geodesics, LLC, Eugene, Oregon). The EEG cap was placed over the top of tACS carbon-silicone electrodes. Data were imported to MATLAB via EEGLAB (40), bandpass filtered (1-40 Hz), down-sampled to 200 Hz, and noisy channels were manually interpolated. Data were re-referenced to average reference. ICA was applied to remove artifacts using ICLabel (41). Power spectral density (PSD) was computed via Welch’s method. The FOOOF toolbox (42) was used to decompose the PSD into periodic and aperiodic components (2-40 Hz, max 3 peaks, fixed aperiodic mode). IAF was defined as the peak within 6-12 Hz from the flattened (aperiodic component removed) PSD.

Alpha PSD was extracted from the aperiodic-removed HD-EEG data at both 10 Hz and daily pre stimulation IAF. The following regions of interest (ROIs) were defined (See Supplementary Methods; Fig S1): LPFC, RPFC, Fz, Cz, P3, and P4. These regions were selected to capture stimulation-relevant activity in areas implicated in depression symptomatology (3,43,44). Each region was defined as a cluster of electrodes and averaged to compute ROI-level power and connectivity metrics. Electrode numbers correspond to the 128-channel HydroCel Geodesic Sensor Net and were mapped to a 91-channel subset (consisting of scalp electrodes on the HydroCel Geodesic Sensor Net) for final analyses.

Functional connectivity was assessed using the weighted phase lag index (wPLI), computed from preprocessed EEG data (aperiodic component retained). wPLI was evaluated at three frequency bands: 10 Hz, individual alpha frequency (IAF), and wPLI at peak wPLI frequency (pf-wPLI; defined as the session-specific frequency within 6-13 Hz at which scalp-averaged wPLI was maximal). For each frequency, wPLI was calculated as the mean connectivity between two regions of interest (ROIs), averaged over all pairwise channel combinations within each ROI pair. The primary ROI of interest was LPFC-RPFC, and additional exploratory ROI connections were also investigated (see Supplementary Methods and Results).

### Statistical analyses

#### Descriptive statistics

Clinical and demographic characteristics are presented descriptively for verum and sham groups. Changes in clinical scores between D1 to D5, and D1 to FUP are presented for scores that showed significant correlations with alpha power. Detailed changes across the timepoints are presented in separate work on the same dataset (35).

#### EEG analysis: power and wPLI Within-day analysis

Two-sample t-tests were conducted for each day (D1-D5) to compare daily power and connectivity (wPLI) changes pre to post stimulation between verum vs. sham groups in pre-defined ROIs. We primarily tested within-day change (post-pre) to assess acute, session-specific effects. For both power and wPLI, p-values were corrected for multiple comparisons using the Benjamini-Hochberg false discovery rate (FDR) procedure, with significance set at p_FDR_ < 0.05. Whole-head, topographical analysis was also conducted using cluster-based permutation testing (45) to examine channel-wise power changes. Spatial adjacency was defined using radius = 4 cm; mean degree ≈ 6-12). Clusters were formed from contiguous electrodes with uncorrected p < 0.05, and cluster-level statistics were obtained by summing t-values within each cluster. Significance was determined via 10,000 random permutations of condition labels, controlling the family-wise error rate (FWER) at α = 0.05 (two-tailed). The resulting t-maps and significant electrode clusters were visualized as scalp topographical maps for condition (verum vs. sham) * time (daily post vs. pre) each day.

The relationship between daily post-pre power and wPLI changes were also examined for each ROI pairing in verum vs. sham groups, and a linear model was fit according to the following equation: ΔFC ∼ Δpower × condition, where condition was coded as a categorical variable with sham as the reference level.

Relationships between daily post-pre changes in power/wPLI (whole brain channel-wise, cluster corrected) and clinical outcomes (HDRS-17, BDI, SHAPS, Q-LES-Q-SF) from D5-D1 or FUP-D1 were examined separately for verum and sham groups using whole-brain topographical analyses. For power, cluster-corrected channel-wise Spearman correlations were performed (using the spatial clusters methods described above). For connectivity, edge-wise ΔwPLI (post-pre) matrices were reconstructed for each day, and Spearman correlations were computed between daily edge-wise ΔwPLI and clinical change scores. Multiple comparisons were controlled using the Benjamini-Hochberg FDR correction at q = 0.05 across edges per day.

#### Cumulative change from D1 analyses

Additionally, we also explored cumulative changes (daily post - D1 pre) to assess changes in power/wPLI relative to baseline using t-tests comparing daily post-stimulation alpha power/wPLI change from D1 pre alpha power/wPLI between verum and sham groups.

#### Entrainment effects: IAF and peak fc frequency

To investigate potential entrainment mechanisms, the relationship between participants’ daily pre-stimulation IAF mismatch from 10 Hz and their corresponding daily post-pre changes in power and connectivity (wPLI) was examined using whole-brain topographical correlations. For power, cluster-corrected channel-wise Spearman correlations were computed between IAF mismatch and power change for each day, and for connectivity, FDR-corrected edge-wise Spearman correlations were computed between IAF mismatch and ΔwPLI for each day (in verum and sham groups separately).

## Results

### Demographics

The sample comprised 20 participants (15 female, 5 male) aged 19-70 years. Mean age did not differ significantly between groups (verum: 39.9 ± 20.9 years; sham: 38.2 ± 15.9 years; Welch’s t(16.8) = 0.20, p = 0.84). Sex distribution was identical across conditions (verum: 8 female, 2 male; sham: 8 female, 2 male; χ^2^(1) = 0.00, p = 1.00). Detailed clinical and demographic details can be found in separate work in the same dataset (35).

### Alpha power dynamics reveal late, session-specific decreases on day four of stimulation

#### Daily within-session alpha power changes across stimulation week

In the verum group, post-pre alpha power showed a progressive reduction across D1-D4, reaching statistical significance on D4 relative to sham. Significant reductions in alpha power at both 10Hz and IAF were found in the verum group compared to sham on D4 of stimulation for all ROIs (central, bilateral frontal and bilateral parietal), demonstrated by two sample t-tests comparing daily pre vs. post change between groups (-91.43% to -150.85% change in verum compared to sham across ROIs, all p<0.05, see Fig. 1C, 1D), where decreases in left and right parietal regions at 10Hz additionally survived FDR correction (p_FWE_ < 0.05; Fig. 1A, 1B). Whole brain condition*time (post-pre) channel-wise cluster permutation analysis demonstrated similar results, with a significant decrease in IAF power at left prefrontal and left parietal regions, as well as a decrease in 10Hz power in left and right parietal regions (cluster corrected, pFWE<0.05). There was no association between daily channel-wise alpha power with IAF mismatch from 10Hz (cluster corrected; all p>0.05). Pre-stimulation IAF remained stable across days in both groups, although the sham group displayed greater variability (Supplementary Figure S3, S4).

**Figure 1.**
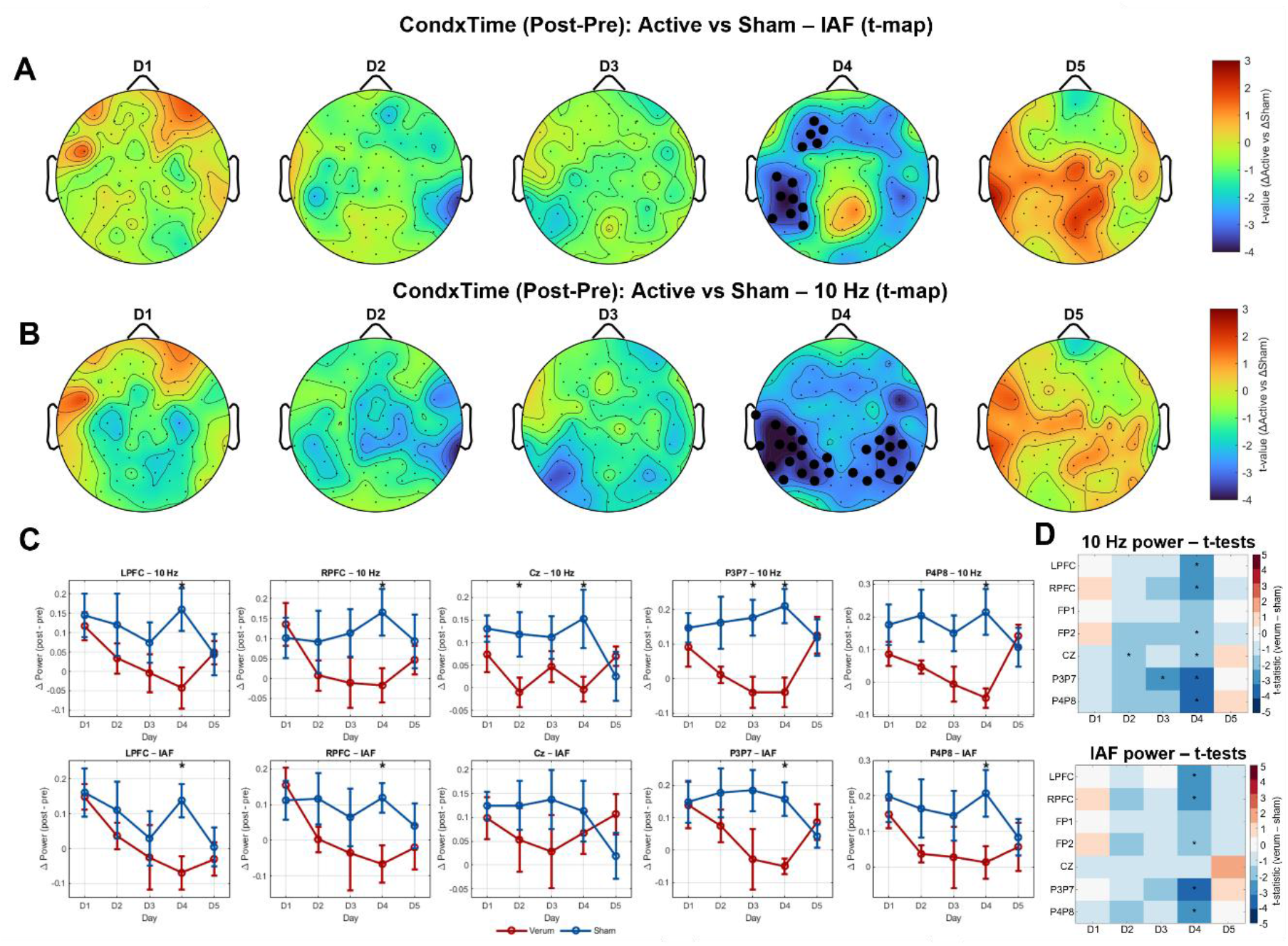
Alpha power is significantly decreased on D4 of stimulation in verum compared to sham group. Trajectory of within-day alpha power changes (post minus pre) in verum and sham groups (aperiodic component removed). Topographical plots of cluster-based permutation tests comparing daily change (post-pre) between verum and sham groups from D1-D5 at (A) IAF and (B) 10Hz. Electrodes marked with solid black dots indicate channels belonging to significant clusters (p < 0.05, cluster-level corrected). Positive t-values reflect stronger power increases (or weaker decreases) in the verum group relative to sham after stimulation, while negative t-values reflect stronger PSD decreases in verum relative to sham after stimulation. Light black dots mark EEG electrodes. (C) Line graphs of daily post minus pre power change in verum (red) and sham (blue) groups across ROIs; error bars indicate standard error of the mean (SEM). (D) Heat maps of IAF and 10Hz power t-statistics for daily pre to post change t-tests between verum and sham groups, indicating significant difference on D4 across most ROIs. Only P3 reductions survived FDR correction for both power metrics (IAF: t=-4.26, p_FDR_ = 0.03; 10 Hz: t=-3.82, p_FDR_ = 0.04). ROI, region of interest; D, day; IAF, individual alpha frequency; LPFC, left prefrontal cortex; RPFC, right prefrontal cortex; PSD, power spectral density;

### Functional connectivity changes occur earlier than power changes, with decreases between days two to three of stimulation followed by an increase by day four

#### Daily within-session wPLI changes across stimulation week

Within day wPLI analysis revealed an early, transient reduction in IAF FC on D2 in the active group across multiple ROI pairs, including bilateral mid-frontal (F3-F4: t=-2.54, p=0.0206), bilateral frontal poles (Fp1-Fp2: t=-2.90, p=0.0094), and right fronto-parietal (F8-P4: t=-2.51, p=0.0220) connections (Supplementary Fig. S1, S5). Further, a significant decrease in F3-Cz 10 Hz wPLI was seen on D4 (t=-2.69, p=0.0150), as well as a trend level sustained decrease in F3-Cz IAF FC across D1-D5 in the verum group (Supplementary Figure S5).

As an exploratory analysis, we next examined cumulative functional connectivity changes, according to daily post-stimulation wPLI change between F3-F4 normalized to baseline. F3-F4 was specifically chosen since these regions were stimulated in-phase during the treatment week, and given the alterations in alpha power and connectivity in left and right prefrontal cortex (46). Results indicated robust cumulative decreases in FC throughout the treatment week, with the strongest effect seen in F3-F4 peak frequency wPLI (Fig. 2). Here, peak-frequency wPLI significantly decreased in the verum vs. sham group on D2 (t=-2.650, p=0.0162) and D3 (D3: t=-2.470, p=0.0237). Smaller, trend level decreases in fc were seen at IAF and 10Hz wPLI at D2 and D3. There were no significant FDR-corrected correlations between daily wPLI and clinical outcomes between D1-D5 or D1-FUP. Finally, there were no significant correlations between daily power and daily wPLI changes.

**Figure 2.**
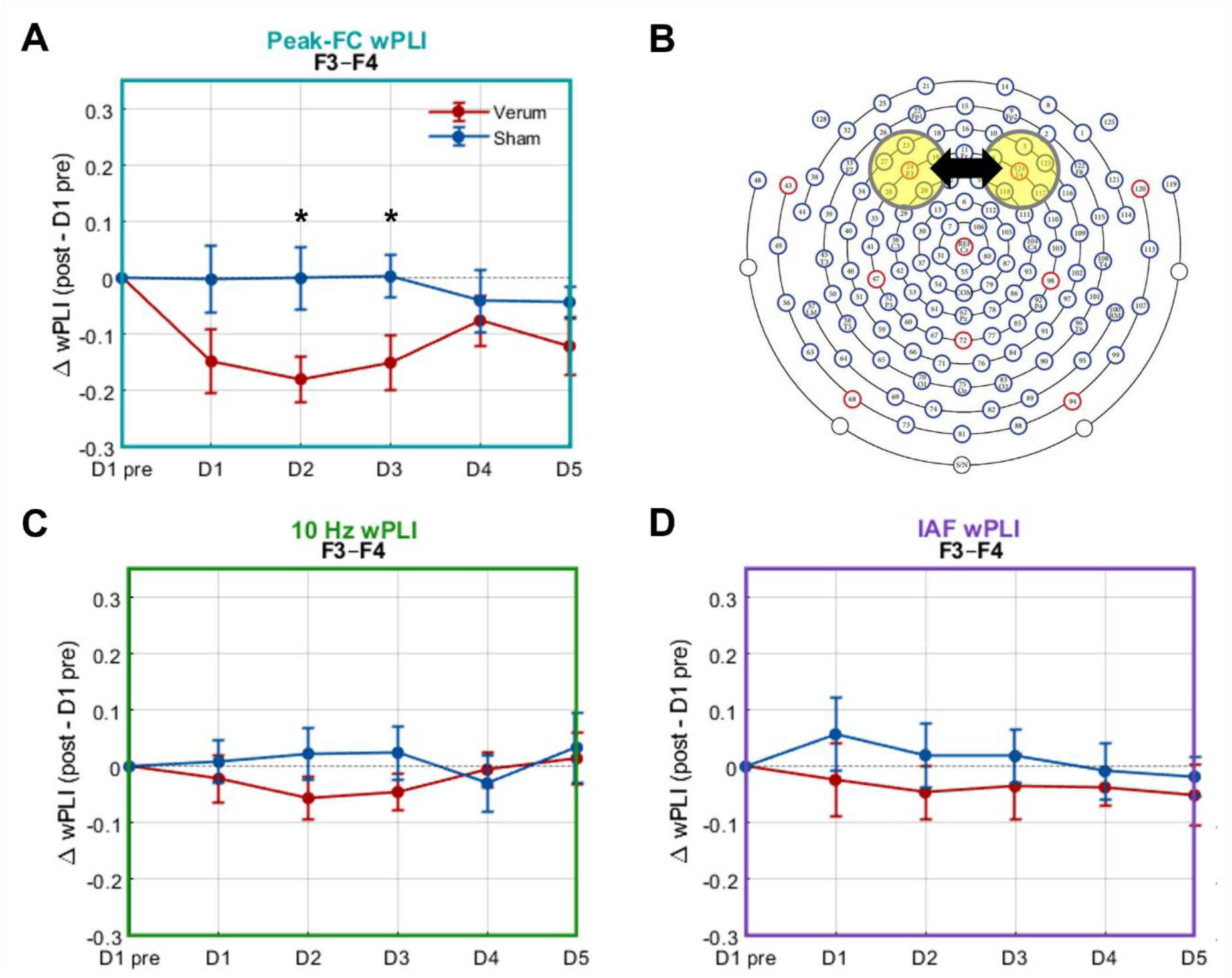
Cumulative functional connectivity decreases between F3-F4 are seen on D2 and D3 relative to baseline at peak FC frequency wPLI only. (A) Peak FC frequency wPLI; (B) Schematic representation of F3-F4 electrode clusters used in wPLI analysis overlain on Geodesic EGI 128 channel EEG net map; (C) 10 Hz wPLI; (D) IAF wPLI. Error bars indicate standard error of the mean.

### Correlations between power and clinical/behavioral outcomes

Next, we explored how immediate within-day changes in power at IAF (whole brain, channel-wise) related to changes in clinical outcomes between D1-D5 or D1-FUP using a whole brain channel-wise approach. There was no significant cluster-wise correlation between change in alpha power on any day and change in depression symptoms at either timepoint.

Further exploratory analysis indicated a significant correlation between D4 IAF power change and change in reward sensitivity (BAS) from D1-D5 (aligning with the significant D4 power decrease), where significantly decreased left prefrontal power was associated with increased reward sensitivity (significant LPFC cluster, spearman’s rho = -0.6, p<0.05) (Fig. 3). This D4 pattern persisted at a trend level for BAS reward sensitivity change from D1 to FUP (Supplementary Figure S6).

**Figure 3.**
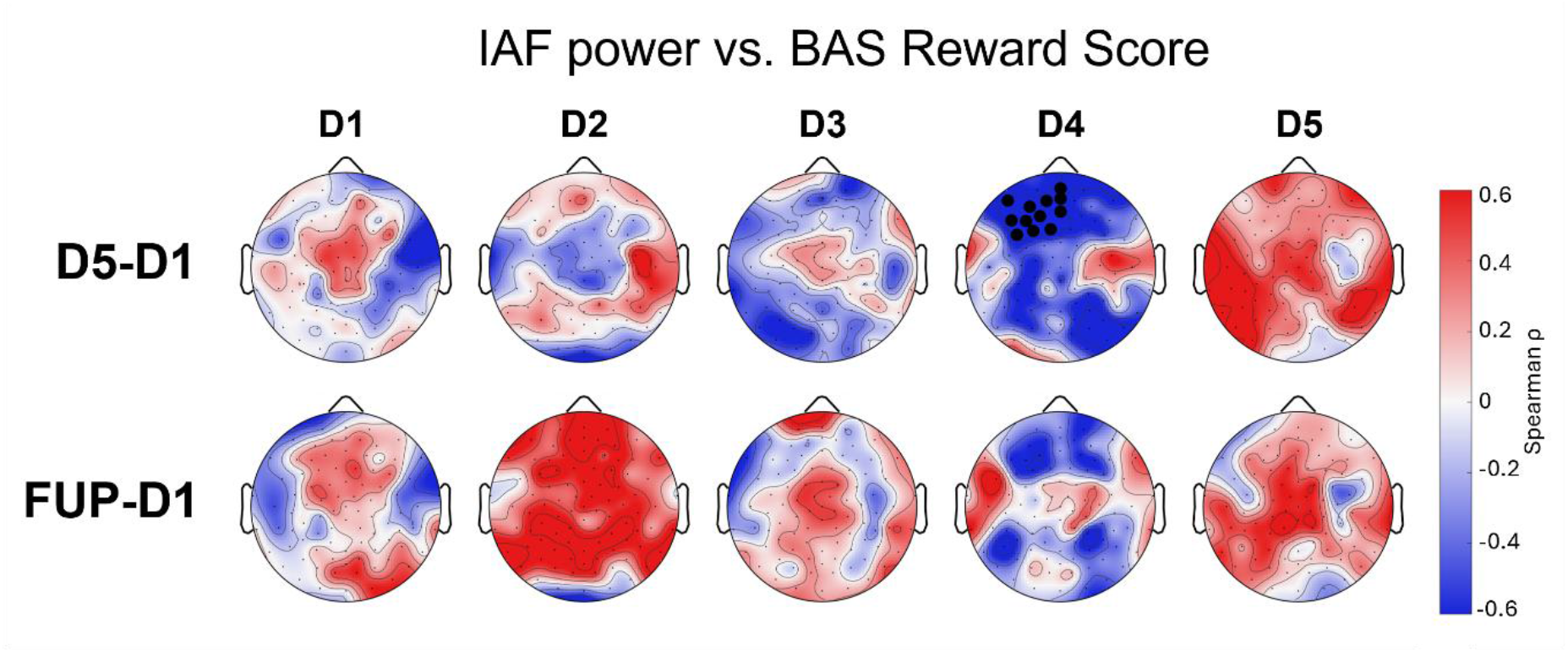
Significant channel-wise correlation between daily within-day IAF power change and BAS drive change (D5-D1 and FUP-D1) in the verum group only. Significant cluster of electrodes in left PFC found for D4 IAF alpha power change vs. BAS reward sensitivity change D5-D1, with similar trend-level patterns at FUP

## Discussion

In this randomized, double-blind, sham-controlled clinical trial of five daily sessions of 10 Hz tACS in MDD, we identified temporally dissociable EEG markers of target engagement: An early, transient reduction in alpha-band functional connectivity between the prefrontal cortices emerged on D2 and D3 relative to D1, followed by a later within-session suppression of alpha power that peaked on D4. These effects were specific to the verum group and were spatially aligned with frontal and fronto-midline networks targeted by stimulation, as well as additional parietal involvement. The D4 within session (post-pre) alpha power suppression in left prefrontal cortex was the only neurophysiological metric that reliably related to clinical or behavioral outcomes, linking left prefrontal alpha suppression to increased reward sensitivity, a construct reflecting heightened responsiveness to positive reinforcement, yet showed no association with depression symptom change. Together, these findings delineate a stage-specific pattern of neural response across repeated tACS sessions and identify D4 alpha suppression as a potential mechanistic marker of behavioral change in a five day tACS trial.

We focused on within-day changes in alpha power to capture the immediate neural response to stimulation. This approach isolates effects that are less visible in cumulative D1-to-D5 measures, providing critical insight to identify when and how target engagement occurs. Across both 10 Hz and IAF metrics, alpha power showed a progressive daily reduction beginning on D2 and reaching a clear maximum decrease on D4, most strongly within left dorsolateral prefrontal and bilateral parietal cortices. This pattern was robust across whole-brain cluster and ROI-based analyses. This left prefrontal alpha power decrease aligns strongly with several previous trials from our group in a single-session IAF stimulation intervention (10), five-day 10Hz stimulation interventions (3,35), and a five-day closed-loop personalized IAF stimulation intervention (6).

Both spatial and temporal patterns of tACS modulation in our study also align with previous work. The spatial topography of the power effects, shown in fronto-parietal rather than purely frontal regions. Frontal stimulation may initially perturb prefrontal circuits, with downstream synchronization changes emerging in parietal regions, which are generally associated with strong expression of alpha oscillations (47,48). This is consistent with a non-linear, multi-phase response trajectory in which early frontal perturbation spreads through large-scale networks, eventually manifesting as broader alpha suppression. Temporally, the D1-D4 pattern aligns with our previous closed-loop IAF findings, where D4 also marked the inflection point linked to clinical improvement (6). Complementing the within-day analysis, in our study, cumulative (i.e., relative to baseline) power changes in the verum group also began diverging from sham on D2, decreasing especially at 10Hz, and continued through D5, consistent with early plasticity mechanisms. The rebound-like increase in D5 alpha paralleled prior work, potentially reflecting a homeostatic response following several days of alpha power suppression (8,43,46,49–51). Together, these findings show a suppression of left frontal and parietal alpha power at both 10Hz and IAF throughout the week, peaking on D4, followed by a trend-level rebound on D5. Our results underscore the underlying complexity of interaction between delivered stimulation and neural response and resulting non-linear effects across multiple consecutive days of stimulation.

Although stimulation was delivered at 10Hz, we examined neurophysiological effects at both 10Hz and IAF, given evidence that tACS is most effective when stimulation matches intrinsic frequency (Arnold Tongue) (30). While power decreases emerged at both frequencies across brain regions, central and parietal alpha power modulation was stronger and earlier at 10Hz: Cz 10Hz power decreased on D3, and cumulative 10Hz reductions emerged by D2. These earlier and broader 10Hz reductions suggest potential entrainment and early spike timing-dependent plasticity at the stimulation frequency (31,52). In contrast, prefrontal alpha reductions were stronger at IAF, shown by both whole-brain cluster and ROI-based effects. IAF remained stable throughout the stimulation week, and we found no modulating effect of the stimulation/endogenous frequency mismatch on daily neural dynamics. Human alpha is not a unitary rhythm but a family of partially independent generators that often organize as anterior-to-posterior traveling waves (48). Engagement of a parietal 10 Hz oscillator early in the stimulation week could reflect the preferential entrainment of posterior alpha sources that naturally exhibit stronger power and more coherent traveling-wave propagation. In contrast, later prefrontal changes at IAF may reflect modulation of a separate, slower anterior alpha generator that is less readily entrained at 10 Hz but is more relevant for depressive pathophysiology.

Interestingly, only the IAF power decrease was associated with behavioral/clinical changes. The D4 reduction in left prefrontal IAF power was moderately to strongly correlated with improved reward sensitivity, with trend-level associations extending to reward sensitivity and goal pursuit at follow-up. This pattern echoes previous work showing that tACS specifically modulates anhedonia symptoms in depression (10,53), and aligns with the central role of prefrontal alpha dynamics in reward processing (54). The absence of clinical correlations at 10 Hz suggests that although the fixed frequency stimulation broadly entrained alpha activity, IAF-specific modulation represented the functionally meaningful neural change. Our previous work demonstrating strong correlations between decreased alpha power and HDRS-17 improvement supports the clinical relevance of IAF effects (6). Together, these findings support future tACS protocols tuned to IAF to maximize both neurophysiological entrainment and clinical impact.

Given evidence that tACS effects can emerge first through phase-relationship shifts rather than changes in oscillatory amplitude (55), we examined both within-day and cumulative functional connectivity throughout the stimulation week. Peak-frequency connectivity between F3-F4 decreased both within D2, as well as cumulatively on D2 and D3 normalized to baseline, preceding the D4 power decrease. This matches earlier work in our group, where five days of 10Hz tACS in people with schizophrenia and schizo-affective disorder resulted in an immediate change in functional connectivity, followed by a more gradual suppression of alpha power (55). The early F3-F4 decoupling suggests that stimulation initially destabilized interhemispheric prefrontal alpha circuits, pushing them out of their baseline synchronous state. We also observed a D4 decrease in 10 Hz F3-Cz connectivity in the verum group, paralleling the D4 power decrease and consistent with an antiphase stimulation montage-driven perturbation of frontal-midline coupling. However, the connectivity-power relationship remains unclear, as changes in wPLI and power did not significantly correlate. Further, connectivity changes were not associated with clinical outcomes. Overall, the connectivity findings point to early engagement of frontal and frontal-midline networks implicated in depression (56,57), including the DMN (58) (59– 61) (62).

A strength of this study was the use of daily high-density EEG to provide greater spatial precision and temporal nuance in neurophysiological measurement, thus permitting quantification of immediate stimulation changes on each day of the intervention. Several methodological considerations should be noted when interpreting these findings. First, the HD-EEG net was placed directly over stimulation electrode pads (F3, F4, Cz). While this design allowed immediate pre and post-stimulation EEG recording, it led to bridged EEG electrodes in these regions, creating a loss of spatial resolution. Second, the sample size was modest, limiting power. Finally, clinical assessments were conducted at D1 and D5, which may not have captured faster-evolving or transient symptom changes that could align with the day-specific neural effects observed here. Future work should incorporate IAF-tuned stimulation and higher-resolution clinical time courses to better capture day-to-day changes.

## Conclusion

Together, these results demonstrate that the neurophysiological effects of five consecutive days of tACS do not reflect a simple cumulative strengthening of stimulation effects, but instead a potential staged sequence of neural processes. These processes may involve early changes in frontal functional connectivity, followed by suppression of left prefrontal alpha activity that peaked on stimulation day four, and related to symptom improvement. Although fixed-frequency 10 Hz stimulation produced widespread entrainment-like effects, only modulation at each individual’s intrinsic alpha frequency related to clinical benefit, highlighting the functional importance of personalized frequency tuning. These findings establish IAF alpha suppression as a promising mechanistic marker of tACS response and point toward stimulation protocols that leverage endogenous oscillations rather than imposing fixed frequencies. Future work optimizing frequency personalization, stimulation timing, and network-level targeting may enhance the therapeutic efficacy of tACS for depression.

## Data Availability

All data produced in the present study are available upon reasonable request to the authors

## Acknowledgements

The authors utilized ChatGPT to assist with language refinement.

## Author Contributions

**Athena Stein:** Methodology, Data Curation, Formal Analysis, Writing – Original Draft, Visualization. **Tobias Schwippel:** Methodology, Data Curation, Investigation, Formal Analysis, Writing – Review & Editing. **Francesca Pupillo:** Data Curation, Investigation. **Hadden LaGarde:** Data Curation, Investigation. **Mengsen Zhang:** Supervision, Writing – Review & Editing. **David Rubinow:** Supervision, Writing – Review & Editing. **Flavio Frohlich:** Conceptualization, Funding Acquisition, Resources, Supervision, Writing – Review & Editing.

All authors: Approved the final version of the manuscript.

## Data Availability Statement

Data from this article will be made available upon reasonable request.

