## Supplementary Material for "Daily EEG reveals stage-specific alpha power and functional connectivity modulation across five days of tACS in major depressive disorder"

### Methods

#### ROI definitions

For power analyses, we defined the following ROIs: LPFC (E19, E20, E23, E24, E27, E28), RPFC (E3, E4, E117, E118, E123, E124), Fz (E10, E11, E16, E18), Cz (E7, E31, E55, E80, E106, E129), P3 (E51, E52, E58, E59), and P4 (E91, E92, E96, E97). For wPLI analyses, we additionally defined the following ROIs: Fp1-Fp2, F3-Cz, F4-Cz, Fz-Pz, F7-P3, F8-P4. ROI electrode clusters were as defined above in the power section, as well as additional Fp1 (E18, E22) and Fp2 (E9, E10) regions.

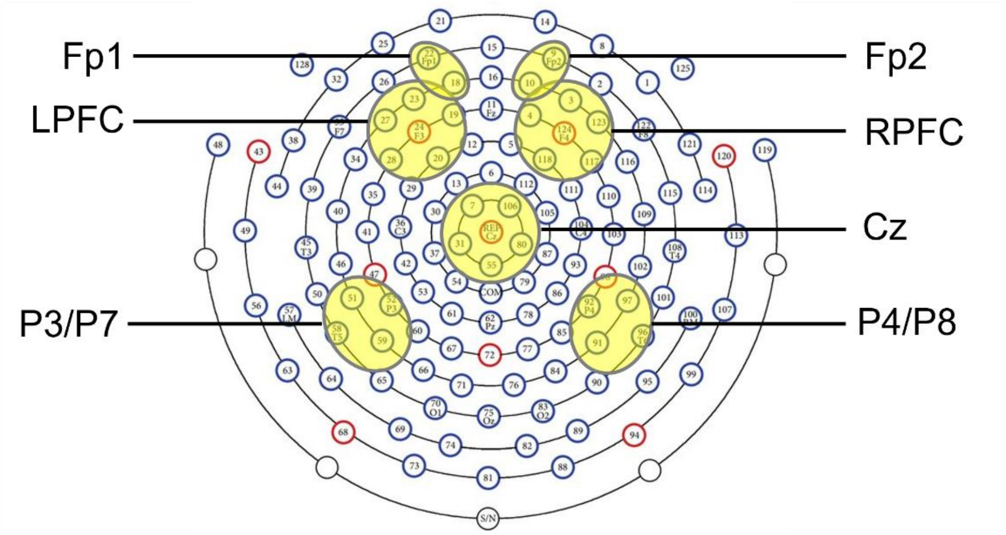


**Figure S1:** Visual representation of each ROI definition overlaid on schematic diagram of 128 channel Geodesic EEG net.

### Results

#### Participants

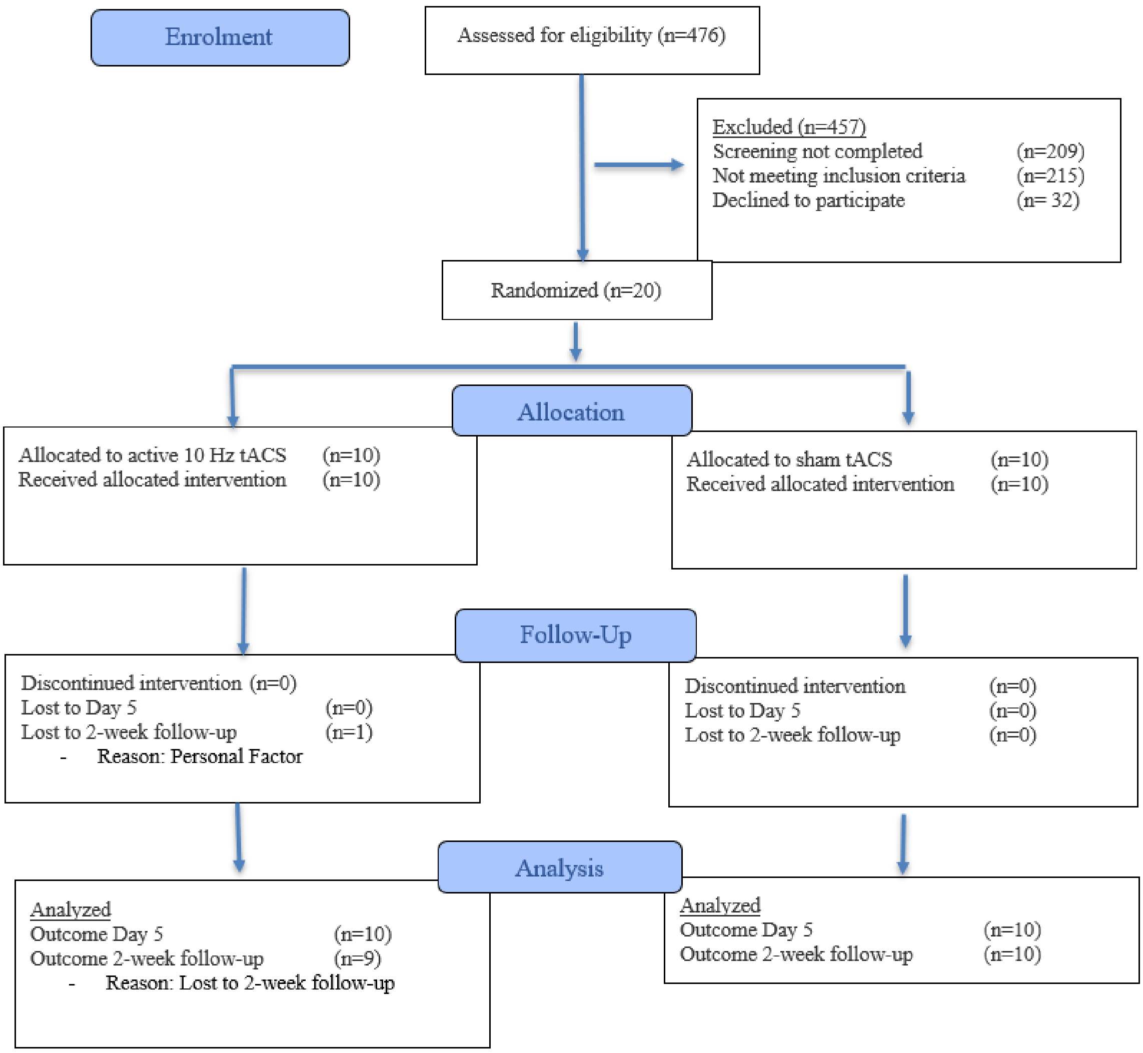


**Figure S2:** CONSORT Diagram.

#### Cumulative Power

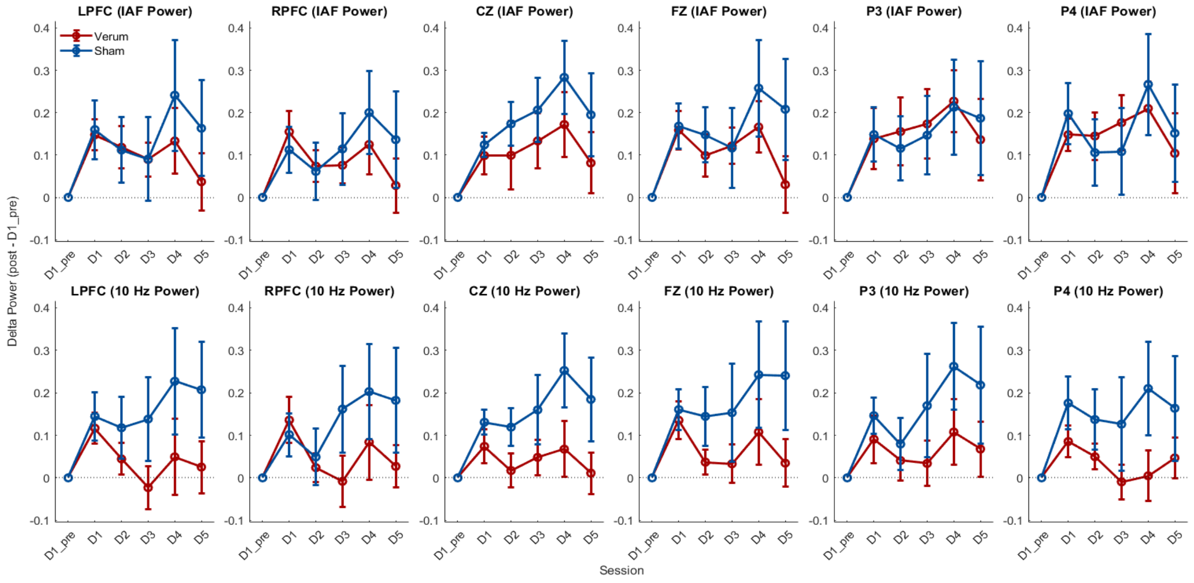


***Figure S3.*** Cumulative trajectory of daily power change (normalized to baseline) for F3, F4, Cz, Fz, P3, P4 ROIs at IAF and 10Hz. Asterisk indicates significance at $p<0.05$.

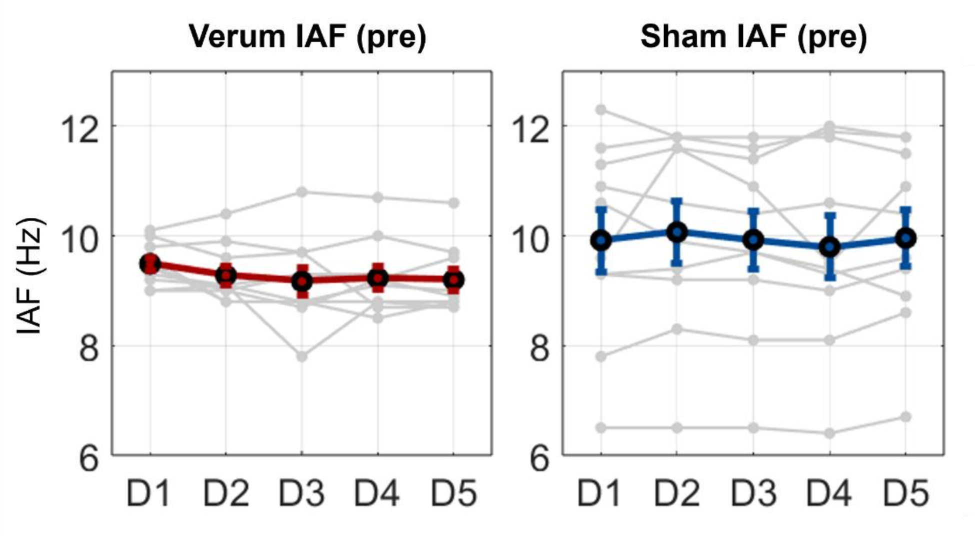


**Figure S4.** *Distribution of daily pre-stimulation IAF in active and sham groups.*

#### Within day functional connectivity for significant ROIs

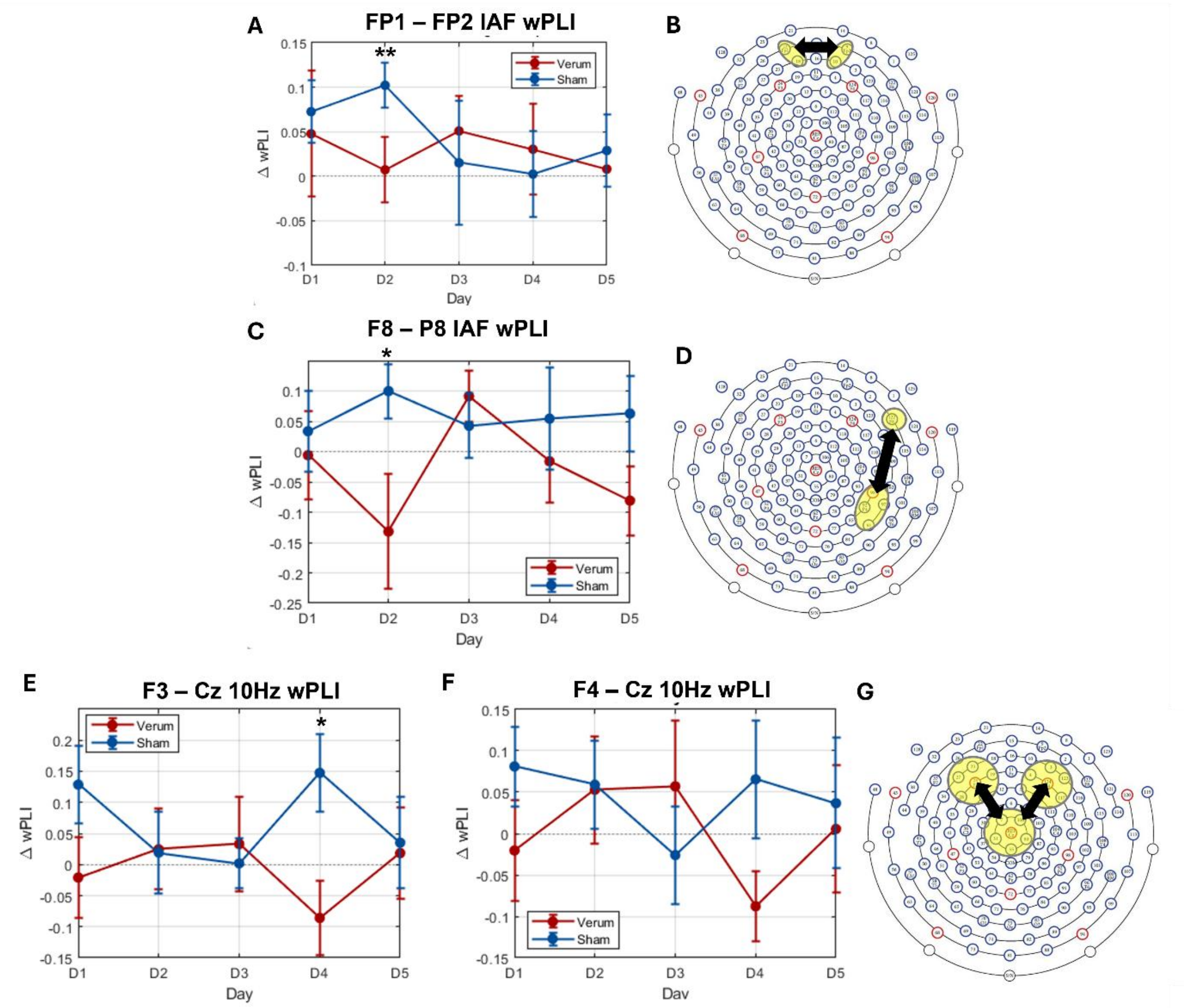


***Figure S5: Functional connectivity decreases are seen on D2 for left-right frontal ROIs at IAF and D4 for left frontal-central ROIs at 10Hz, complementing power results.*** *(A,C,E,F) Line graphs indicating decreased pre-post wPLI in verum group compared to sham on D2 (IAF) and D4 (10Hz), * indicates significance at p<0.05, ** indicates significance at p<0.001. (B,D,G) Schematic representation of electrode clusters used in wPLI analysis overlain on Geodesic EGI 128 channel EEG net map.*

**Power vs. clinical scores: topographic correlations**

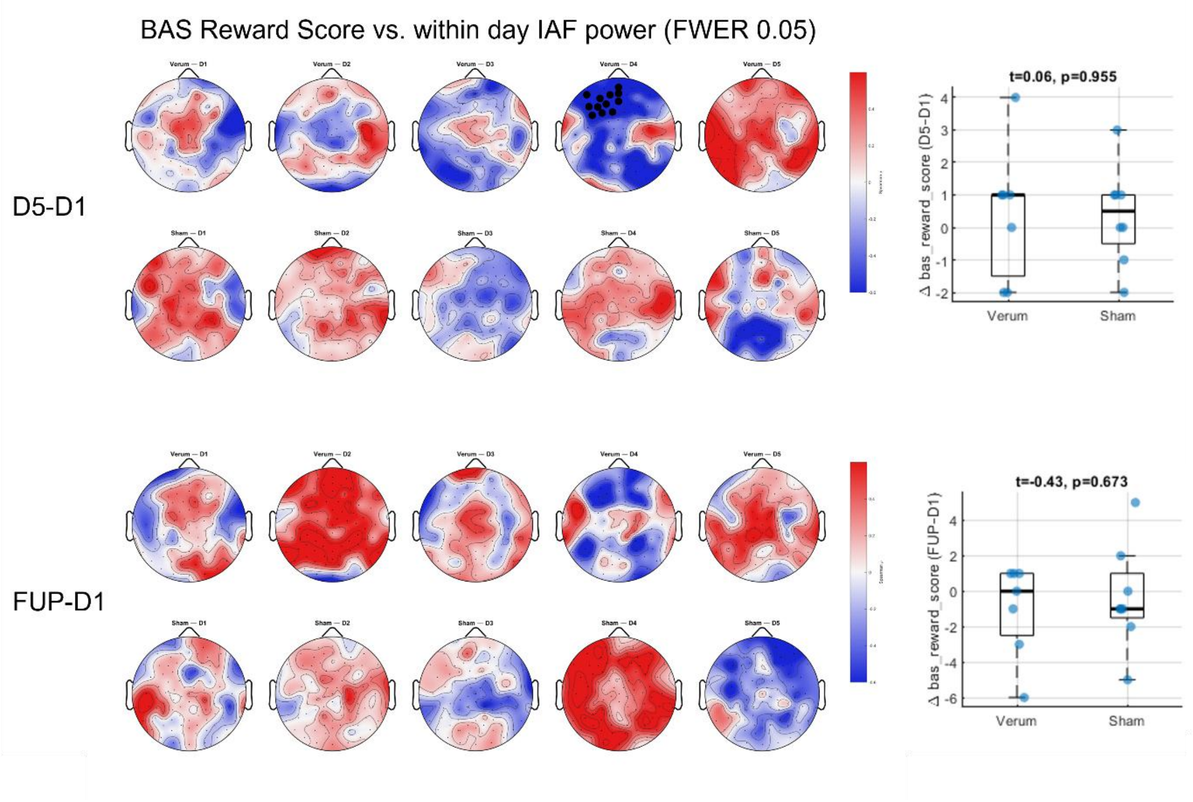


**Fig. S6: Significant electrode-wise correlation between within day IAF power and change in BAS reward sensitivity (D5-D1 and FUP-D1).** Boxplots indicate distribution of clinical score change in active and sham groups
